# Exploring Perspectives of Healthcare Staff on Achieving the Healthcare Quality Standards in District Hospitals in Rwanda: A qualitative Study

**DOI:** 10.64898/2026.11.07.26367515

**Authors:** Jovial Thomas Ntawukuriryayo, Gillian Woolhead

## Abstract

**Background:** The World Health Organization (WHO) has recommended that quality of care (QoC) should be timely delivered within safe conditions based on patients’ needs and preferences. However, low- and middle-income countries face challenges such as understaffing and inadequate infrastructure, resulting in poor QoC. In 2012, Rwanda leveraged the pre-existing performance-based financing (PBF) to introduce continuous quality improvement (QI) towards accreditation recognition of district hospitals (DH). Despite these efforts, no DHs accredited by 2022.

**Objective:** To explore healthcare professionals’ (HCP) perspectives on achieving the delivery of standard QoC in Rwandan DHs.

**Methods:** Using an interpretative epistemology, this qualitative study conducted in-depth semi-structured interviews with 14 participants purposively sampled HCPs from Masaka (urban) and Remera-Rukoma (rural) DHs. Interview data were audio-recorded, manually transcribed, and analyzed with thematic content analysis.

**Results:** The analysis revealed three main themes: perceptions about continuous QI towards accreditation recognition, barriers to achieving accreditation, and strategies to achieve the recognition. Participants from both hospitals expressed that the accreditation program required more efforts than the PBF approach. They also highlighted that the DHs were still at the awareness stage and had not clearly implemented the quality standards due to challenges such as low staff involvement and motivation, staff turnover and shortage, and limited budget for accreditation. Recommended strategies to achieve the recognition included staff motivation through increased incentives and salaries, cascaded accreditation rollout, integrating accreditation program into pre-service curriculum.

**Conclusions:** This qualitative study highlighted the challenges, such as limited budget and low staff motivation, impeding progress towards DH accreditation recognition. Cascaded accreditation rollout, enhanced staff incentives and salaries, and integration of accreditation program into pre-service curriculum were key strategies suggested for Rwanda Ministry of Health and DHs to achieve the required quality standards and recognition.

## Introduction

Quality standards of healthcare services are critical for achieving the best patient’s health outcomes through the delivery of effective, safe, and timely evidence-based interventions based on patients’ needs and preferences within the same quality standards across all patients^1,2^. However, low quality care―including, but not limited to, inaccurate diagnosis, unnecessary or inappropriate treatment, and medication errors―can lead to increased morbidity and mortality, reduced trust in health systems, and substantive wasted expenditures for health systems and patients^3,4^.

In low- and middle-income countries (LMICs), low quality of care is responsible for eight million deaths per year, causing a loss in the economic welfare of $6 trillion^3^. Healthcare providers in these countries often deliver less than 50% of the recommended evidence-based healthcare and around one third of patients experience poor communication, disrespectful care, long waiting times, and short consultation. Less than 25% of people in the LMICs are satisfied with care services provided in their healthcare systems as opposed to 50% of people in high-income countries (HICs)^3^.

In Rwanda, quality of care in hospitals also faced patient safety issues. For instance, in 2019, 10-20% of treatment delivered to patients was not needed for specific diseases, due to inaccurate diagnoses^5^. In addition to existing high workload and limited staffing levels (Rwanda Public Health Bulletin, 2024), other examples were the commonly reported incidents which included medication errors (25.3%), loss to follow-up (25%), and surgical site infections (20%)^5^. In 2016, patient waiting time for seven hospitals varied from three hours 32 minutes to seven hours 33 minutes across all hospital services. By 2019, this waiting time was still high with some hospitals having over six hours^6^, reflecting poor delivery of timely care^7^. Additionally, at least 75% of health professionals were dissatisfied with their salaries and benefits (Rwanda Public Health Bulletin, 2024), reflecting that providing quality healthcare by generally dissatisfied staff is difficult.

In response to the challenge of low quality of care, Rwanda aligned healthcare services with quality standards by leveraging the pre-existing performance-based financing (PBF) system and initiated continuous quality improvement (QI) towards accreditation recognition in all district hospital^8^. This program was implemented five risk areas including leadership process and accountability, competent and capable workforce, safe environment for staff and patients, clinical care, and improvement of quality and safety^9^. Each of these domains has a set of quality standards and each standard is organised into three levels of effort. Level one focuses on the availability of written organisational policies and clinical protocols as well as staff awareness of the documents. Level two is about the implementation of what is described in the guiding documents and level three focuses on data-driven decision-making to continuously improve quality of healthcare^8,9^.

However, since 2012, none of the hospitals have yet received accreditation recognition for providing healthcare services meeting quality standards, reflecting ongoing challenges of achieving the standard quality of care. In addition, while healthcare staff working in the hospitals are on the frontline providing quality health services, there was a need for improving their willingness to implement quality standards along with their routine activities among their top priorities^5^, which can help with maximising healthcare effectiveness, timeliness, and patient safety^7^.

The most recent assessment conducted in 44 district hospitals―including those that were recently upgraded to Level Two Teaching Hospitals, referral hospitals, and Provincial Hospitals―found that 61% of the hospitals were recognized for having met the requirements for level I and II, and 36% of the hospitals for level I only, while achieving a recognition for level III remains a significant challenge^10^.

As the healthcare professionals are a critical workforce in the active implementation of the quality standards, exploring their perspectives^11^ on how to overcome the challenges and achieve the quality standards is essential for a successful system redesign and implementation of measures for accelerating the attainment of the standard quality of care in the district hospitals in Rwanda. This study therefore aimed to explore healthcare professionals’ perspectives on how to achieve standard quality of care services in district hospitals, in order to inform enhancement of the healthcare quality in Rwanda.

## Methods

### (a) Research paradigm and researcher reflexivity

This study used the constructivist and interpretivist approaches^11,12^ to explore views of healthcare professionals about appropriate strategies for achieving the standard quality of healthcare.

As the researchers could be seen as part of the research participants and then considered as an ‘insider’^13^, the investigator critically reflected on values and beliefs that could influence the research findings^14^. The interviewer allowed the participants to express their thoughts based on their own experience and knowledge. Additionally, the interviewer ‘bracketed’ their views to mitigate any background impact, used a reflexive diary throughout the research process by assuring participants that there were no correct answers^15^.

### (b) Design and Setting

This study used a qualitative study design with in-depth semi-structured interviews to obtain rich information from the participants. This design was chosen because it enables to gain deeper insights into people’s experiences and allows the opportunity to explore people’s views, beliefs, and perceptions in depth^16^. The study was conducted at two district hospitals, Masaka Hospital in Kigali city and Remera-Rukoma in rural area.

### (c) Recruitment

Purposive sampling was used by selecting different types of healthcare professionals that included medical doctors, nurses/midwives, allied health professionals, and supervisors of health services in order to ensure information rich cases are selected so as to enable a deeper understanding of the study subject^17^. Participants were recruited through snowball sampling through recommendation of previous participants^18^. Participants were included in the study if they were over the age of 18, worked at the district hospital, were familiar with and had been actively involved in the healthcare accreditation activities.

### (d) Data Collection

We conducted 14 qualitative individual in-depth interviews with the healthcare professionals for 30-45 minutes each, from January 04 to February 08, 2022.

To test recruitment procedures, comprehensiveness of the interview guide questions and if they were collecting the right concepts that answer the research question^19^, we conducted a pilot testing with the first two participants who fulfilled the inclusion criteria. The interview guide was not revised because the pilot results showed that the questions were comprehensive and collected the right data. The pilot testing participants were then included in the data analysis as there were no changes made to the interview guide.

The interview was recorded with a smartphone sound recorder and supplemented with notes taken at each interview. The researcher aimed to reach data saturation which is the point in the research when no new information is discovered in the data analysis^20^.

### (e) Data Analysis

The interviews were transcribed verbatim and translated the transcripts into English, where necessary. A thematic content analysis was used to uncover themes, sub-themes, and emerging concepts^21^. The codes that described a specific pattern were organised into a broader theme. The reviewed patterns and themes were reorganised into codes, subthemes, and themes, and presented in a codebook (Table 1).

**Table 1.** Codebook.

| THEMES | SUBTHEMES/CATEGORIES | CODES |
|---|---|---|
| Perception about quality improvement towards accreditation recognition | Accreditation standards | Beginning of the accreditation process |
|  | Support for accreditation programme | Hospital-generated revenues |
|  | Accreditation status | Progress towards the accreditation recognition |
|  | Estimated time of achieving accreditation recognition | Time to achieve accreditation recognition |
| Barriers to achieving accreditation recognition | Limited Financial resources | Insufficient financial resources |
|  |  | No donors and partners specific to accreditation |
|  | Low compliance with policies/protocol | Low staff motivation |
|  |  | Inadequate provision of PBF |
|  |  | Staff turnover/ shortage |
|  |  | Same salaries for different degrees and responsibilities |
|  |  | Low staff involvement |
|  |  | Inability to comply with standards due to limited equipment and infrastructure |
|  |  | Many responsibilities |
|  |  | Accreditation is in words, but not in practice |
|  | Low leadership involvement | Weak/decreasing efforts by hospital leaders |
| Recommended strategies to address the barriers to achieving accreditation recognition | Staff motivation and commitment | Salary/incentive increase for staff |
|  | Increased leadership involvement | Cascaded accreditation rollout |
|  |  | Integrate QI and accreditation into pre-service curriculum |
|  | Staff training | Staff training and awareness raising |
|  |  | Vigorous and continuous supervision |
|  |  | Losing interest in maintaining accreditation recognition |
|  |  | Achieving accreditation recognition by cheating |

#### Ethical Considerations

All participants confirmed their participation in the study by signing their informed consent. This research was approved by the University of Liverpool (ETH-101121-MPH256), Rwanda National Health Research Committee (NHRC/2021/PROT/041), Rwanda National Ethics Committee (836/RNEC/2021), and authorized by Rwanda Ministry of Health (20/8035/DPMEHF/2021). Informed consent― in which confidentiality and anonymity were guaranteed―was sought and obtained from each participant before data collection.

## Results

### (a) Perception about quality improvement towards accreditation recognition Accreditation standards

Almost half of the participants from both urban and rural facilities said that before the continuous quality improvement (QI) towards accreditation of district hospitals (DH), the staff were used to the pre-existing performance-based financing (PBF) approach which was much easier than the accreditation process as staff could prepare for the PBF assessment just before it started and get good performance scores. The hospitals were first given some standards known as “patient safety goals” of quality improvement (QI) in combination with the PBF system and thereafter, started the full accreditation process.

> *“…. we previously had a PBF system, where the scoring and evaluation systems were not tough like in accreditation; they used to check the completeness of patients’ medical files, and these files were completed in the last days or one month just before the evaluation; people could get good scores … in districts, the accreditation started as patient safety goals … thereafter, we were given all the standards …”* (I1).

### Support for accreditation programme

Many participants highlighted that donors and partners such as Ingobyi Activity supported some QI activities, including equipment provision and staff training in maternal and child health programme, but not directly to the accreditation programme. The Ministry of Health (MOH) also provided accreditation facilitation and biomedical equipment to all the DHs regardless of geographic location. The local district administration also supported the DHs in quality of care (QoC) in many ways including analysis of patients’ suggestions and complaints and provision of a report with recommendations to the hospital administration. Using a portion of financial resources from their own revenues from healthcare services delivered to patients, the hospitals included a budget line for accreditation into their annual plan and budget. A few participants indicated that the budget allocated to accreditation activities was small but did not estimate its percentage based on the overall budget.

### Accreditation status

Overall, the status of accreditation within the DHs was still at the awareness stage or level one across both rural and urban settings. The implementation was done, but it was not yet secured as mentioned by most of the participants:

> *“… the current status, I think, we are still at the awareness stage. I can’t say that the implementation is clear, we are still dealing with awareness, we are still learning the policies and procedures, plans; we also do implementation, but it is not yet at 100% looking at the current status …”* (I1).

### Estimated time of achieving accreditation recognition

The healthcare professionals had different views about when the hospitals would be recognised, with the shortest time predicted being two years from the time of conducting this study and the longest period being 20 years. Some participants said it would take a long time to be accredited, but it was not easy for them to predict when their hospital would achieve the recognition as there were many challenges to address. Other participants felt the question was very hard to answer and preferred not to answer it as the existing challenges could not allow them to predict at all.

> *“… (laughter), uh! That question is difficult! It’s like asking me when Jesus will come. If we had capacities, enough staff, money … it cannot take time to be accredited* (I10).

### (b) Barriers to achieving accreditation recognition

#### Limited financial resources

The hospitals were struggling with implementing the accreditation activities due to a limited budget. Most of the participants said that the hospitals used a small portion of their own revenues to prepare only for the external accreditation assessment. This challenge made it difficult for them to comply with the standards within the existing budget constraints as stated by many participants especially those from rural areas.

> *“… the limited budget restricts us to comply with standards … The hospital does not have enough resources for covering the cost of most of the things …”* (I10).

#### Low compliance with policies/protocols

Participants said that there was low compliance with the quality standards due to high workload, staff shortage and turnover, inadequate infrastructure and equipment, and low salaries and financial (PBF) incentives. The staff turnover and shortage limited the ability to deliver quality care to patients and required the DHs to usually assign accreditation responsibilities to staff who already had other responsibilities. For instance, risk area two could be given to the Human Resource Manager, who was already overburdened with daily HR management activities and ended up not finding time for the accreditation duties. The QI Officer therefore had to work on all the accreditation activities alone as said by almost half of the participants from both facilities. In addition, the incentives, which was expected to increase with improved accreditation performance, did not change upon achieving better performance, demotivating the staff. Linking this incentive with the hospital’s performance demotivated staff. For instance, if a hospital did not provide the required facilities for effective hand hygiene and marked low during an assessment, the staff ended up receiving a reduced PBF amount. Most of the participants from both urban and rural DHs said that it was not clear how the staff were penalised for organisational responsibilities.

> *“… By the time of the accreditation, people were promised a top-up upon achieving a level. But achieving level one, and even level two, they did not receive the promised incentives. This affected the performance, we lost level two on the following assessment … people reduced their commitment in the accreditation activities … there are many things to improve …”* (I13).

Most of the participants from both settings highlighted that the low financial motivation was the leading factor for staff turnover. The high workload, low salaries and PBF incentives were behind the low motivation for the existing staff, which also triggered resistance to accreditation requirements for some staff. However, other staff were involved under the leadership’s influence, but they immediately relaxed upon leadership change, especially when the efforts of new leaders were weak.

Additionally, a few participants emphasised that due to the existing challenges across the urban and rural DHs, the staff used to present false documents to surveyors during the assessment, which implied that the accreditation process was more in words than in practice; reflecting the hospitals’ inability to maintain achieved successes.

> *“… our barriers across the district hospitals are similar. Today we succeed, tomorrow we fail … a hospital is unable to stabilise the implementation. It seems as if the accreditation is in words, but not in practice … I can say that all of this leads us, during the accreditation assessment, to present things, false documents …”* (I14).

#### Low leadership involvement

Some hospital leaders might not put effort into the accreditation activities, which decreased performance in some risk areas, discouraging other staff who work hard to maximise their performance. Some participants noted that the poor performance that was frequently seen in risk areas one and two, which are mainly for the leaders, was behind the leadership’s weakness.

> *“… If the leadership is sleeping, we also, we sleep … if I miss people who are always there to encourage me, I end up disregarding the accreditation … If the risk area 4 has the highest score, but the lowest scores are reported in the administrative risk areas … what does it mean? The leadership doesn’t care about that …”*. (I9).

### (c) Recommended strategies to address the barriers to achieving accreditation recognition

Recommendations for addressing barriers to accreditation of healthcare quality mainly focused on aligning salaries and financial incentives for healthcare professionals current market prices and enhancing leadership involvement in cascaded rollout of the healthcare quality programme and integrating the healthcare quality accreditation into pre-service curriculum.

#### Staff motivation and commitment

Most of the participants recommended the MOH harmonise PBF incentives and salaries across hospitals according to the living standards to ensure that staff are not moving from one health facility to another.

> *“… if the leaders can increase financial motivation for accreditation levels …they achieve this level and you provide this amount, it can help to maximise the performance. And for the staff turnover, it wouldn’t be there as people are already motivated …”* (I12).

#### Increased leadership involvement

Many participants believed that the MOH needed to consider the use of a cascaded approach for the accreditation rollout. For instance, the process could start with a cohort of hospitals until they are fully accredited, support another cohort up to accreditation recognition, and continue the same process until all the hospitals are accredited.

> *So, in my opinion, I suggest one thing, the Ministry of Health can identify like 5 district hospitals, 2 provincial hospitals, and 1-2 referral hospitals, and support those hospitals in the accreditation process up to level three, and after having these fully accredited, then focus on supporting another group of hospitals; but lifting all the hospitals as a whole, it becomes like stressful, you end up finding that no one is progressing. I don’t support the idea of lifting all the district hospitals to level three. No. Those who have not achieved level one, should focus on achieving that level and then move to the next level instead of just moving on to level three, without being stable at previous levels. Imagine a student who fails on a simple exam, can that student succeed on a more difficult exam? He/she can’t. That’s another challenge that discourages the hospitals*.

Most participants from both rural and urban settings also emphasised that the MOH could consider integrating accreditation standards into pre-service curriculum for students in health-related fields, for them to start practicing in the hospitals when they already know what accreditation is and what is expected from them.

> *Another suggestion is to deliver the accreditation course to all students who will potentially come across accreditation, who will work in healthcare facilities, physicians, nurses, laboratory, all of them; particularly in district hospitals. In that way the students would come into hospitals when they are already aware of what accreditation is and what is expected from them*.

## Discussion

This study explored barriers to the accreditation process across the district hospitals (DH) and the strategies needed to speed up the process towards the accreditation recognition. The participants perceived the continuous quality improvement (QI) towards accreditation recognition as a difficult implementation process because of different challenges. Additionally, the fact that the DHs were still struggling with improving the awareness of the process, the participants were generally unsure of the expected timeline for achieving the accreditation recognition.

The barriers to the progress towards achieving this recognition as highlighted by most of the participants included limited funding sources for the continuous QI activities as the DHs did not have external support directly related to accreditation. Instead, these hospitals needed to find a budget from their own revenues which were even insufficient for addressing existing operational problems. Other studies similarly found that the high cost of implementing accreditation standards was one of the great barriers to effective implementation, which requires sufficient quality equipment and adequate health infrastructure^22,23^. Another barrier was low staff motivation due to low staff incentives and salaries, leading to low compliance with the standards. Due to the low compared to the cost of living, the hospital staff had to leave the hospital to work for other institutions where they received more incentives and higher salaries. This turnover created an additional workload on the remaining staff with some staff having multiple responsibilities, and finally contributed to a decline in the performance of QI activities within the DHs. This result supports other qualitative studies, where it was found that the staff shortage and turnover were key barriers to the implementation of hospital accreditation programmes and a hospital tends to lose its best performing employees when it is unable to motivate them^23,24^.

This study found that the DH staff sometimes presented incorrect documents during the accreditation assessment. This issue was mainly due to the existing challenges including the low motivation, high workload, insufficient facilitation, and limited equipment and materials which restricted the staff to comply with standards as required. As the accreditation evaluation was also linked to the PBF incentives, the staff presented false documents to surveyors just to please their leaders and receive the incentives. Besides, the behaviour of presenting incorrect documents to surveyors might have been a custom as some participants mentioned that staff used to complete documents just before the PBF evaluation. This practice implies that during the accreditation assessments, the staff continued to present things that were not in their routine practice. A qualitative study conducted to explore challenges of hospital accreditation model in Iran also found that hospital staff used to present false documentation to surveyors, mainly due to existing challenges including lack of incentives and staff shortage^25^.

This study also found that cascading the implementation of the accreditation programme across the DHs could be a good approach to speed up achieving the DH accreditation recognition. With this strategy, the MOH could focus efforts on one cohort of DHs up to the accreditation recognition and progressively continue with other cohorts until all DHs are accredited. Rwanda had initially introduced the full accreditation programme with five DHs, with the target of helping them to achieve level one after two years. However, after four of the five hospitals achieved level one, the programme was immediately extended to all the DHs^8^, which the participants thought was too heavy for the national health system.

Most participants also believed that integrating the hospital accreditation programme into pre-service curriculum would facilitate fresh graduates to start clinical practice in the DHs when they are already aware of what accreditation is and required quality expectations. This strategy aligns with the WHO recommendation of integrating QI and assurance into the pre-service curriculum for health professional programmes^7^.

## Strengths and Limitations

To ensure the trustworthiness of these research findings, this study used different strategies including verbatim transcription to show that the data were collected from the research participants^26^. The credibility of the research findings was reflected in the fact that the investigator had a previous experience in healthcare quality. This opportunity allowed him to ask pertinent questions to get a deeper understanding of the participants’ insights^27^. To ensure dependability, the details of the findings were clearly described, allowing repeatability of the work^28^. As the data collection reached saturation, the results can be transferred to other DHs^26^. However, it may not be feasible to use the findings in other health facilities including private and national referral hospitals as the scope of work is different. The confirmability of the findings reflected in the fact that participants were asked to express their thoughts freely as there were no correct or wrong answers^28,29^. However, due to the COVID-19 pandemic restrictions, the investigator conducted telephone interviews as face-to-face data collection was not possible. The researcher could also take note of non-verbal information such as facial expressions through video call interviews, but all the participants did not prefer to use any video call platforms.

## Conclusion

The district hospitals (DH) in Rwanda had been running a hospital accreditation programme since 2012. However, until 2022 there were not even one of the hospitals accredited for providing standard quality of care. The research participants from both rural and urban DHs highlighted major barriers to achieving accreditation recognition, including low staff involvement, staff turnover and shortage, and inadequate infrastructure and equipment. The recommended strategies for achieving and maintaining the recognition included cascaded accreditation rollout, financial staff motivation, leadership involvement and commitment through cascaded rollout and integration of accreditation programme into pre-service curriculum.

## Data Availability

All data produced in the present study are available upon reasonable request to the authors.

## Acknowledgements

This study was conducted as part of academic requirement for the Master of Public Health at the University of Liverpool. Acknowledgements are directed to the university, and particularly, the Commonwealth Scholarship Commission for funding my studies.

## Author Contributions

JTN: Study conceptualisation, data collection, data analysis, and manuscript writing.

GW: Manuscript review and supervision.

## Conflict of Interest

The authors have no conflict of interest to declare.

## Notes

### Competing Interest Statement

The authors have declared no competing interest.

## References

1. European Commission. So What? Strategies across Europe to Assess Quality of Care. European Union; 2016. doi:10.2875/89450

2. WHO. Quality Health Services. World Health Organization. July 20, 2020. https://www.who.int/news-room/fact-sheets/detail/quality-health-services

3. Kruk M, Gage A, Joseph N, Danaei G, Garcia-Saiso S, Salomon J. Mortality due to low-quality health systems in the universal health coverage era: a systematic analysis of amenable deaths in 137 countries. Lancet Glob Health. 2018;392(10160):2203–2212. https://www.thelancet.com/journals/lancet/article/PIIS0140-6736(18)31668-4/fulltext

4. WHO. Improving the quality of health services: tools and resources. Turning recommendations into practice Geneva: World Health Organization. Preprint posted online 2018.

5. Ndagijimana D, Mureithi C, Ngomi NN. Quality and Safety Management of Health Care Service Delivery among Public Hospitals in Rwanda: a Cross-Sectional Survey. InternatIonal Journal of TranslatIonal MedIcal Research and PublIc Health. 2019;3(2):95–106. doi:10.21106/ijtmrph.81

6. Achuri D, Wilson DR, Atwine J, Kamuhangire E. Reducing Patient Waiting Times in Rwandan Hospital Outpatient Services. 2019. Accessed July 12, 2021. https://www.msh.org/sites/default/files/cfa_technical_highlight_for_eop_final_to_print.pdf

7. WHO, World Bank Group, OECD. Delivering Quality Health Services. 2018. https://extranet.who.int/sph/sites/default/files/document-library/document/9789241513906-eng.pdf

8. Binagwaho A, Scott KW, Dushime T, et al. Creating a pathway for public hospital accreditation in Rwanda: progress, challenges and lessons learned. International Journal for Quality in Health Care. 2020;32(1):76–79. https://academic.oup.com/intqhc/article/32/1/76/5536100

9. Rwanda MOH. Rwanda Hospital Accreditation Standards. 2022.

10. Rwanda MOH. Accreditation Performance Progress Report 2020/2021. 2021.

11. Soini H, Kronqvist E, Huber G. Epistemologies for Qualitative Research. Center for Qualitative Psychology. 2011;8. http://www.aquad.de/nexus/cqp-nexus-08.pdf

12. Pulla V, Carter E. Employing Interpretivism in Social Work Research. International Journal of Social Work and Human Services Practice. 2018;6(1):9–14. Accessed June 8, 2021. https://www.researchgate.net/publication/340920288_Employing_Interpretivism_in_Social_Work_Research/fulltext/5ea42ba1a6fdccd79451e251/Employing-Interpretivism-in-Social-Work-Research.pdf

13. Kerstetter K. Insider, Outsider, or Somewhere Between: The Impact of Researchers’ Identities on the Community-Based Research Process. J Rural Soc Sci. 2012;27(2). Accessed May 20, 2021. https://egrove.olemiss.edu/cgi/viewcontent.cgi?article=1434&context=jrss

14. Dixon-Woods M, Campbell A, Chang T, et al. A qualitative study of design stakeholders’ views of developing and implementing a registry-based learning health system. Implementation Science. 2020;15(1). doi:10.1186/S13012-020-0976-1

15. Tufford L, Newman P. Bracketing in Qualitative Research: Qualitative Social Work. Published online 2010. doi:10.1177/1473325010368316

16. Busetto L, Wick W, Gumbinger C. How to use and assess qualitative research methods. Neurol Res Pract. y2020;2(14). doi:10.1186/S42466-020-00059-Z

17. Gentles S, Charles C, Ploeg J, McKibbon K. Sampling in Qualitative Research: Insights from an Overview of the Methods Literature. The Qualitative Report. 2015;20(11):1772–1789. http://www.miguelangelmartinez.net/IMG/pdf/2015_Gentles_Sampling_Qualitative_ResearchTQR.pdf

18. Naderifar M, Goli H, Ghaljaie F. Snowball Sampling: A Purposeful Method of Sampling in Qualitative Research. Strides in Development of Medical Education. 2017;14(3). doi:10.5812/sdme.67670

19. Majid MAA, Othman M, Mohamad SF, Lim SAH, Yusof A. Piloting for Interviews in Qualitative Research: Operationalization and Lessons Learnt. International Journal of Academic Research in Business and Social Sciences. 2017;7(4). doi:10.6007/IJARBSS/v7-i4/2916

20. Saunders B, Sim J, Kingstone T, et al. Saturation in qualitative research: exploring its conceptualization and operationalization. Qual Quant. 2018;52(4):1893–1907. doi:10.1007/s11135-017-0574-8

21. Vaismoradi M, Jones J, Turunen H, Snelgrove S. Theme development in qualitative content analysis and thematic analysis. Journal Nursing Education and Practice. 2016;6(5). doi:10.5430/jnep.v6n5p100

22. Yousefinezhadi T, Mosadeghrad AM, Hinchcliff R, Akbari-Sari A. Evaluation results of national hospital accreditation program in Iran: The view of hospital managers. J Healthc Qual Res. 2020;35(1):12–18. doi:10.1016/J.JHQR.2019.08.008

23. Algunmeeyn A, El-Dahiyat F, Alfayoumi I, Alrawashdeh M. Exploring staff perspectives of the barriers to the implementation of accreditation in Jordanian hospitals: Case study. Int J Healthc Manag. 2021;14(4):1422–1428. doi:10.1080/20479700.2020.1763233

24. Alshamsi AI, Thomson L, Santos A. What Impact Does Accreditation Have on Workplaces? A Qualitative Study to Explore the Perceptions of Healthcare Professionals About the Process of Accreditation. Front Psychol. 2020;11. doi:10.3389/FPSYG.2020.01614/FULL

25. Vali L, Mehrolhasani MH, Mirzaei S, Oroomiei N. Challenges of implementing the accreditation model in military and university hospitals in Iran: a qualitative study. 2020;20:698. doi:10.1186/s12913-020-05536-4

26. Houghton C, Casey D, Shaw D, Murphy K. Rigour in qualitative case-study research. Nurse Res. 2013;20(4):12–17. http://pdfs.semanticscholar.org/65fa/d576b17038113098b53864ed3aa08af70aee.pdf

27. Ritchie J, Lewis J, Nicholls C, Ormston R. Qualitative Research Practice: A Guide for Social Science Students & Researchers. 2nd edn. SAGE; 2014. https://elearning-uol.vitalsource.com/#/books/9781446293515/cfi/6/2!/4/2/2@0:36.5

28. Johnson JL, Adkins D, Chauvin S. A Review of the Quality Indicators of Rigor in Qualitative Research. Am J Pharm Educ. 2020;84(1):138–146. doi:10.5688/AJPE7120

29. Cypress BS. Rigor or reliability and validity in qualitative research: Perspectives, strategies, reconceptualization, and recommendations. Dimensions of Critical Care Nursing. 2017;36(4):253–263. doi:10.1097/DCC.0000000000000253

